# Mediterranean Diet and atherothrombosis biomarkers: a randomized controlled trial

**DOI:** 10.1101/19001909

**Authors:** Álvaro Hernáez, Olga Castañer, Anna Tresserra-Rimbau, Xavier Pintó, Montserrat Fitó, Rosa Casas, Miguel Ángel Martínez-González, Dolores Corella, Jordi Salas-Salvadó, José Lapetra, Enrique Gómez-Gracia, Fernando Arós, Miquel Fiol, Lluis Serra-Majem, Emilio Ros, Ramón Estruch

## Abstract

**Scope:** To assess whether following a Mediterranean diet (MedDiet) improves atherothrombosis biomarkers in high cardiovascular risk individuals.

**Methods and results:** In 358 random volunteers from the PREDIMED trial (*Prevención con Dieta Mediterránea*), we assessed the 1-year effects on atherothrombosis markers of an intervention with MedDiet, enriched with virgin olive oil (MedDiet-VOO; *N*=120) or nuts (MedDiet-Nuts; *N*=119) versus a low-fat control diet (*N*=119). In a secondary, observational approach, we studied whether volunteers with large increments in MedDiet adherence (>2 score points) were associated with 1-year improvements in biomarkers (relative to those worsening their adherence). The MedDiet-VOO intervention increased platelet activating factor-acetylhydrolase activity in high-density lipoproteins (HDLs) by 7.5% [95% confidence interval: 0.17; 14.8] and decreased HDL-bound α_1_-antitrypsin levels by 6.1% [−11.8; −0.29]. The MedDiet-Nuts one reduced non-esterified fatty acid concentrations by 9.3% [−18.1; −0.53]. Only the low-fat diet was associated with increases in platelet factor-4 and prothrombin factor_1+2_ levels versus baseline (*P*=0.012 and *P*=0.003, respectively, according to Wilcoxon signed-rank tests). Finally, large MedDiet increments were associated with less fibrinogen (−9.5% [−18.3; −0.60]) and non-esterified fatty acid concentrations (−16.7% [−31.7; −1.74]).

**Conclusion:** Following a MedDiet improves atherothrombosis biomarkers in high cardiovascular risk individuals.

## INTRODUCTION

Better conformity with a traditional Mediterranean Diet (MedDiet) prevents major cardiovascular clinical outcomes^[1–3]^. These beneficial effects may be mediated by improvements in risk factors related to glucose metabolism, endothelial function, lipid profile, oxidative stress, and low-grade inflammation^[4]^. Nevertheless, little evidence related to MedDiet effects on atherothrombosis mechanisms is available, although the antithrombotic properties of some of its individual components are known^[5,6]^. Increased levels of biomarkers related to platelet aggregation (P-selectin^[7]^, platelet factor-4^[8]^) and coagulation (fibrinogen^[9]^, prothrombin fragment 1+2 –proportional to thrombin formation– ^[10]^), and decreased concentrations of fibrinolysis indicators (plasminogen activator inhibitor-1 –PAI-1–^[11]^, D-dimer^[12]^) are linked to an increased risk of suffering atherosclerotic events. In addition, high levels of non-esterified fatty acids (NEFAs) and dysfunctional high-density lipoproteins (HDLs) have also been attributed pro-thrombotic properties^[13,14]^. In the context of the PREDIMED (*Prevención con Dieta Mediterránea)* trial, following a MedDiet decreased P-selectin levels^[15]^. Beyond this study, only a small-scale prospective analysis with 21 young, healthy male volunteers indicated that following a MedDiet-like dietary pattern was related to lower fibrinogen levels and an attenuation of the coagulation response^[16]^, and two cross-sectional studies have reported associations between MedDiet adherence and less D-dimer and fibrinogen values^[17,18]^. However, no intervention trial has studied the long-term effects of this healthy dietary pattern on a comprehensive set of biomarkers of atherothrombosis to date.

Our main aim was to assess whether a 1-year intervention with MedDiet improved a set of atherothrombosis biomarkers in high cardiovascular risk individuals. Our secondary aim was to determine whether 1-year changes in MedDiet adherence were associated with 1-year differences in these indicators.

## EXPERIMENTAL SECTION

### Study design

Study subjects were participants of the PREDIMED Study. It was a large-scale, parallel, multicenter, randomized controlled trial assessing the long-term effects of following a MedDiet on the primary prevention of cardiovascular disease in high cardiovascular risk individuals^[1]^. Volunteers were men (aged 55-80) and women (aged 60-80) free of cardiovascular disease with type 2 diabetes mellitus or at least three of these factors: high levels of low-density lipoprotein cholesterol, low HDL cholesterol concentrations, hypertension, overweight/obesity, current smoking, or family history of premature heart disease. Further details of inclusion/exclusion criteria are available in **Supplemental Methods**. The trial protocol complied with the Declaration of Helsinki, was approved by local institutional ethic committees, registered under the International Standard Randomized Controlled Trial Number ISRCTN35739639 (http://www.isrctn.com/ISRCTN35739639), and described in previous publications^[1,19]^. An institutional ethic committee (CEIC-PSMAR) also approved the protocol of the present study. All participants provided written informed consent before joining the study. Individuals were randomly assigned to one of three interventions (in a 1:1:1 ratio): 1) a MedDiet enriched with virgin olive oil (MedDiet-VOO); 2) a MedDiet enriched with mixed nuts (MedDiet-Nuts); and 3) a low-fat control diet. MedDiet interventions promoted: 1) the consumption of fruits, vegetables, pulses, nuts, and fish; 2) the use of virgin olive oil as main culinary fat; 3) a decrease in the intake of ultra-processed foods/drinks and animal fats and the substitution of red/processed meats for poultry; and 4) the consumption of foods cooked by home-made methods (such as the traditional “sofrito”). To promote compliance and account for family needs, volunteers allocated in the MedDiet-VOO intervention received 1L/week of virgin olive oil and those in the MedDiet-Nuts group 210 g/week of mixed nuts. Volunteers allocated to the low-fat control group (following the 2002 recommendations of the American Heart Association) were advised: 1) to promote the intake of fruits, vegetables, and pulses; 2) to decrease the consumption of high-fat dairy products, butter/margarine, meat, and ultra-processed foods/drinks, and 3) to moderate their intake of other fatty foods (olive oil, nuts, and fatty fish). Further details of the dietary intervention are available in **Supplemental Methods**.

For the present analyses, we selected a random subsample of 358 subjects (4.8% of the total PREDIMED population) recruited in Hospital Clinic (Barcelona) and Hospital del Mar Medical Research Institute (Barcelona) study sites, with fasting citrate plasma samples collected at baseline and after 1 year of intervention. 120 volunteers were allocated to the MedDiet-VOO intervention group, 119 to MedDiet-Nuts, and 119 to the control diet. Samples were stored at −80°C until the analyses. The CONSORT checklist regarding our study is available in **Supplemental Table 1**.

**Table 1.** Description of the study population.

|  | MedDiet-VOO<br>(*n*=120) | MedDiet-Nuts<br>(*n*=119) | Low-fat diet<br>(*n*=119) | *P*-value |
| --- | --- | --- | --- | --- |
| Age (years) (mean, SD) | 67.3 (5.34) | 66.6 (5.79) | 66.5 (6.35) | 0.530 |
| Sex (female) (*n*, %) | 78 (65.0%) | 75 (63.0%) | 74 (62.2%) | 0.898 |
| Educational level: |  |  |  | 0.228 |
| Primary/less (*n*, %) | 95 (79.2%) | 79 (66.4%) | 81 (68.1%) |  |
| Secondary (*n*, %) | 15 (12.5%) | 25 (21.0%) | 21 (17.6%) |  |
| Higher (*n*, %) | 9 (7.50%) | 14 (11.8%) | 13 (10.9%) |  |
| Non-available (*n*, %) | 1 (0.83%) | 1 (0.84%) | 4 (3.36%) |  |
| Type-2 diabetes mellitus (*n*, %) | 67 (55.8%) | 55 (46.2%) | 61 (51.3%) | 0.331 |
| Hypercholesterolemia (*n*, %) | 88 (73.3%) | 86 (72.3%) | 90 (75.6%) | 0.834 |
| Hypertension (*n*, %) | 95 (79.2%) | 92 (77.3%) | 91 (76.5%) | 0.877 |
| Use of antithrombotic drugs (*n*, %) | 19 (15.8%) | 13 (10.9%) | 14 (11.8%) | 0.479 |
| Tobacco use: |  |  |  | 0.445 |
| Never smoker (*n*, %) | 83 (69.2%) | 77 (64.7%) | 69 (58.0%) |  |
| Actual smoker (*n*, %) | 16 (13.3%) | 15 (12.6%) | 20 (16.8%) |  |
| Former smoker (*n*, %) | 21 (17.5%) | 27 (22.7%) | 30 (25.2%) |  |
| Weight status |  |  |  | 0.717 |
| Normal weight (*n*, %) | 8 (6.67%) | 4 (3.36%) | 7 (5.88%) |  |
| Overweight (*n*, %) | 56 (46.7%) | 62 (52.1%) | 62 (52.1%) |  |
| Obesity (*n*, %) | 56 (46.7%) | 53 (44.5%) | 50 (42.0%) |  |
| Mediterranean diet adherence score<br>(mean, SD) | 8.61 (1.68) | 8.69 (1.85) | 8.99 (1.64) | 0.197 |
| Leisure-time physical activity<br>(metabolic equivalents of task·min/w)<br>(median [1<sup>st</sup> quartile; 3<sup>rd</sup> quartile]) | 1,505<br>[724; 2,566] | 1,253<br>[538; 2,345] | 1,410<br>[416; 2,639] | 0.447 |
| Alcohol intake (g/week)<br>(median [1<sup>st</sup> quartile; 3<sup>rd</sup> quartile]) | 12.1<br>[0.00; 53.6] | 30.7<br>[0.00; 94.3] | 10.2<br>[0.00; 79.7] | 0.175 |

*MedDiet-Nuts*: Mediterranean diet enriched with mixed nuts; *MedDiet-VOO*: Mediterranean diet enriched with virgin olive oil.

### Mediterranean diet adherence and covariates

MedDiet adherence was estimated at baseline and after 1 year of intervention using the MedDiet adherence score. It was a validated short screener interrogating whether the volunteer followed the essential 14 dietary characteristics associated with a MedDiet (positively scoring: the intake of virgin olive oil, nuts, fruits, vegetables, legumes, fish, and wine in moderation; and the substitution or avoidance of animal fats, red and processed meats, processed foods, and sugary drinks)^[20]^. 1-year increments in MedDiet adherence were calculated by subtracting the baseline adherence to the 1-year value. Data on age, sex, educational level, body mass index, smoking habit, and presence of type-2 diabetes mellitus, hypercholesterolemia, hypertension, leisure-time physical activity, and use of antithrombotic medications were gathered at baseline by trained clinical personnel^[1,19]^.

### Atherothrombosis biomarkers

We used ELISA kits to quantify the levels of: fibrinogen (*Human Fibrinogen SimpleStep ELISA kit*, ref.: ab208036, abcam, UK), PAI-1 (*Human PAI1 SimpleStep ELISA kit (SERPINE1)*, ref.: ab184863, abcam, UK), platelet factor-4 (*PF4 (CXCL4) Human SimpleStep ELISA kit*, ref.:ab189573, abcam, UK), and prothrombin fragment 1+2 (*Human F1+2 (prothrombin fragment 1+2) ELISA kit*, ref.: E-EL-H1793, Elabscience, USA). We assessed the concentrations of antithrombin (*Antithrombin III*, ref.: SR-1102016, Spinreact, Spain) and D-dimer (*D-Dimer*, ref.: SR-1709231, Spinreact, Spain) by immunoturbidimetry, and of NEFAs (*Non-Esterified Fatty Acids*, ref.: FA115, Randox, Spain) using a colorimetric technique in an ABX-Pentra 400 autoanalyzer (Horiba-ABX, France). All the previous analyses were performed in fasting citrate plasma samples. In parallel, in apolipoprotein B-depleted plasma samples (specimen in which all lipoproteins but HDLs were eliminated, prepared from fasting citrate plasma as well^[21]^), we quantified total cholesterol by colorimetry (*Cholesterol Enzymatic-Colorimetry*, ref.: SR-41022, Spinreact, Spain) and α_1_-antitrypsin (*a-1-antitrypsin*, ref.: SR-1102054, Spinreact, Spain) by immunoturbidimetry in an ABX-Pentra 400 autoanalyzer (Horiba-ABX, France). With this data, we calculated the α_1_-antitrypsin/cholesterol ratio (“α_1_-antitrypsin levels in HDL”). We also determined in these samples the activity of the PAF-AH enzyme by a colorimetric kit (*PAF Acetylhydrolase Activity Assay Kit*, ref.: K765-1000, Cayman Chemical, USA). A detailed explanation of the quality control is available in **Supplemental Methods**.

### Sample size

We assumed that the variability of the measurement of P-selectin in a previous PREDIMED sub-analysis would be similar to that in our variables. A sample size of 119 individuals per group (total sample size: *N*=357) allowed ≥80% power to detect a 10% change relative to the mean baseline value of P-selectin levels (7.65 ng/mL) between post- and pre-intervention values, and a 14% change relative to the mean baseline value (10.8 ng/mL) in the differences among the three diets. We considered a 2-sided type I error of 0.05, a loss rate of 5%, and the standard deviation of the differences in P-selectin values in this intervention (SD=28.9 ng/mL)^[15]^.

### Statistical analyses

We first checked the distribution of continuous variables in normality plots and by the Shapiro-Wilk test. We assessed whether there were differences in baseline values among the three intervention groups with one-way ANOVA tests for normally distributed continuous variables, Kruskal-Wallis tests for non-normally distributed continuous ones, and chi-squared tests for categorical parameters.

As main analysis (based on randomization), we evaluated whether there were differences in the 1-year changes in atherothrombosis biomarkers (calculated by subtracting baseline from post-intervention values) in the MedDiet interventions relative to the control diet by multivariate linear regressions. We studied these differences in the whole population of the study (main approach), in subjects with very low adherence to a MedDiet at baseline (<8 score points), and only in the individuals presenting the main cardiovascular risk factors affecting atherothrombosis according to bibliography (type 2 diabetes mellitus, smokers, users of antithrombotic drugs, hypertensive, and hypercholesterolemic)^[22]^. We also analyzed if there were differences between pre- and post-intervention values in every study arm by paired t tests in normally distributed variables and Wilcoxon signed-rank tests in non-normally distributed ones.

As secondary, observational analyses, we studied whether substantial increments in MedDiet adherence after 1 year of intervention (≥3 score points) were associated with 1-year changes in atherothrombosis biomarkers (in relation to decreases in MedDiet adherence) by multivariate linear regression models.

To help in data interpretation, we present the results as percentage changes relative to baseline values. To calculate them, we divided the adjusted difference coefficients obtained in the regression models by the baseline value of the variable in each of the groups (in the main analyses) or in the 358 volunteers (in the secondary analyses). Models were adjusted for: age (continuous), sex, study site, educational level (primary education or lower/secondary or equivalent/higher education), baseline value of the atherothrombosis biomarker, diabetes (yes/no), hypercholesterolemia (yes/no), hypertension (yes/no), use of antithrombotic medication (yes/no), tobacco use (never/former/actual smoker), body mass index (continuous), leisure-time physical activity (continuous), alcohol use (continuous), and two propensity scores to correct for the theoretical deviations in the randomization process (calculated from 30 baseline variables)^[1]^. Secondary, observational analyses were additionally adjusted for the allocated intervention group and MedDiet adherence score at baseline. Models were plotted using the “lme” package in R Software^[23]^.

We performed statistical analyses with R software version 3.5.0^[24]^.

## RESULTS

### Study population and intervention

We found no clinical differences in baseline characteristics among intervention groups (**Table 1**). Relative to the non-included PREDIMED Study population, there were 6.2% more women, 7.2% more hypercholesterolemic individuals, and 7.4% less antithrombotic drug users among our volunteers, and they presented higher physical activity levels (+22 metabolic equivalents of task·min/d) and alcohol intake (+0.67 g/d) (**Supplemental Table 2**).

Volunteers’ compliance improved during the year of intervention (MedDiet-VOO: +1.14 points [95% confidence interval: 0.79; 1.49]; MedDiet-Nuts: +1.35 [1.00; 1.69]). No changes in physical activity were observed (**Supplemental Table 3**).

### Randomization-based analyses: effects of 1-year MedDiet interventions on atherothrombosis biomarkers

In the whole study population, the MedDiet-VOO intervention increased the activity of platelet activating factor-in HDLs by 7.5% [0.17; 14.8] (**Figure 1I**) and decreased HDL-bound α_1_-antitrypsin levels by 6.1% [−11.8; −0.29] (**Figure 1H)**. The MedDiet-Nuts one reduced non-esterified fatty acid concentrations by 9.3% [−18.1; −0.53] (**Figure 1G**).

**Figure 1.**
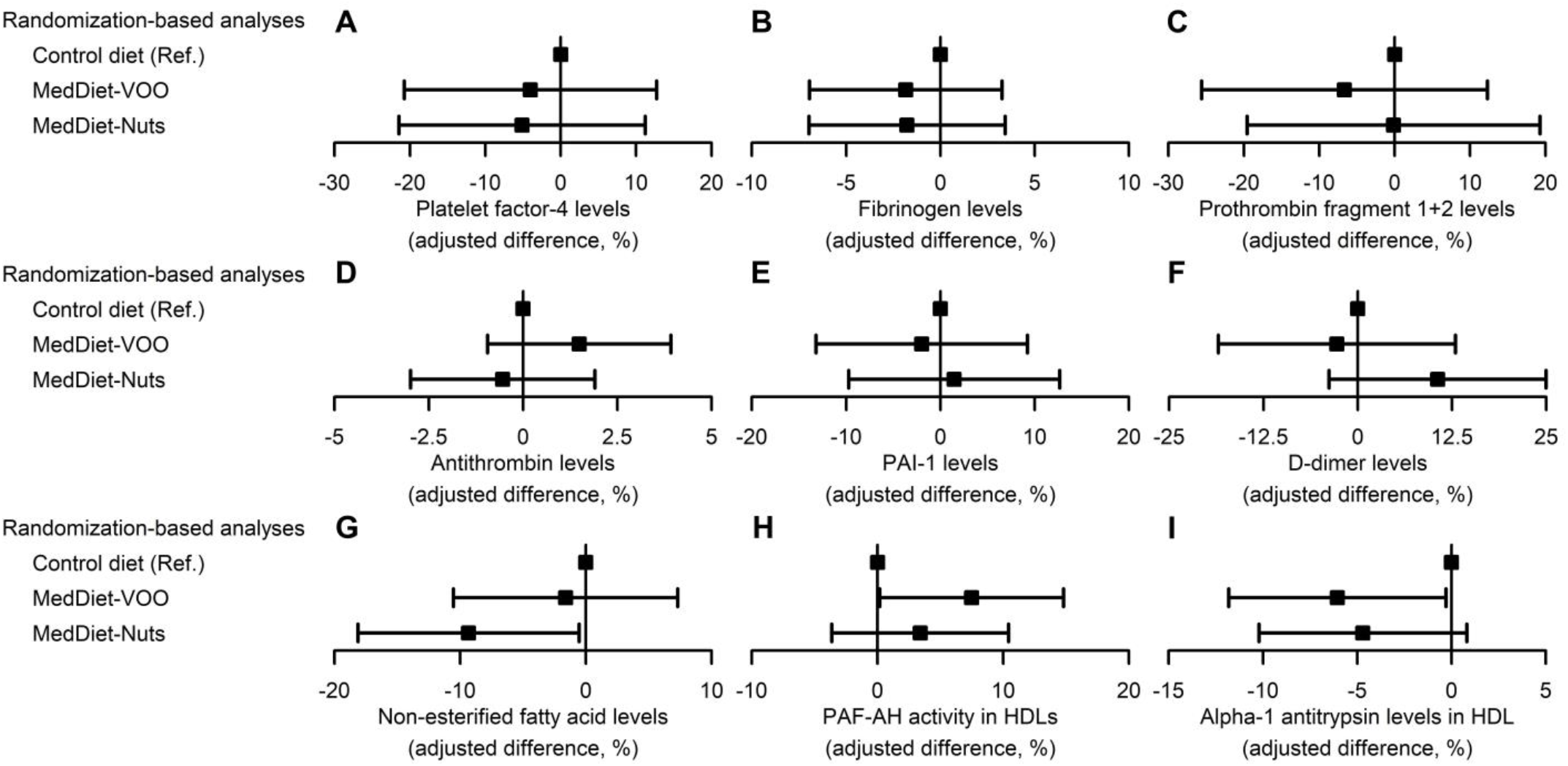
Effects of Mediterranean diet interventions on atherothrombosis biomarkers. Results are shown as adjusted coefficients (percentage changes relative to baseline values) with 95% confidence intervals. *HDL*: high-density lipoprotein; *MedDiet-Nuts*: traditional Mediterranean diet enriched with nuts; *MedDiet-VOO*: traditional Mediterranean diet enriched with virgin olive oil; *PAF*-*AH*: platelet activating factor acetylhydrolase; *PAI-1*: plasminogen activator inhibitor 1.

Volunteers with low MedDiet adherence at baseline and users of antithrombotic drugs particularly benefited from MedDiet interventions regarding some biomarkers in which we did not observe significant changes in the whole study population. Volunteers with low MedDiet adherence particularly increased their antithrombin levels after the MedDiet-VOO intervention (+6.1% [0.76; 11.4]; **Supplemental Figure 1D**). Users of antithrombotic drugs particularly decreased their D-dimer levels after the MedDiet-VOO intervention (−54.1% [−86.8; −21.3]) and a beneficial effect is suggested after the MedDiet-Nuts one (−26.7% [−56.1; 2.78]; **Supplemental Figure 1F**). A decrease in PAI-1 concentrations (−32.3 [−66.5; 2.01]; **Supplemental Figure 1E**) and an increase in antithrombin levels (+8.2% [−1.08; 17.4]; **Supplemental Figure 1D**) are also suggested after the MedDiet-VOO intervention in this population.

When assessing post-vs. pre-intervention changes, only the low-fat control diet was associated with increases in platelet factor-4 concentrations (*P*=0.012) and prothrombin fragment 1+2 levels (*P*=0.003) relative to baseline (**Supplemental Table 4**).

### Secondary analyses: associations of 1-year changes in MedDiet adherence with 1-year differences in atherothrombosis biomarkers

Substantial increments in MedDiet adherence, relative to decreases in compliance, were associated with reduced fibrinogen (−9.5% [−18.3; −0.60]; **Figure 2B**) and NEFA concentrations (−16.7% [−31.7; −1.74]; **Figure 2G**).

**Figure 2.**
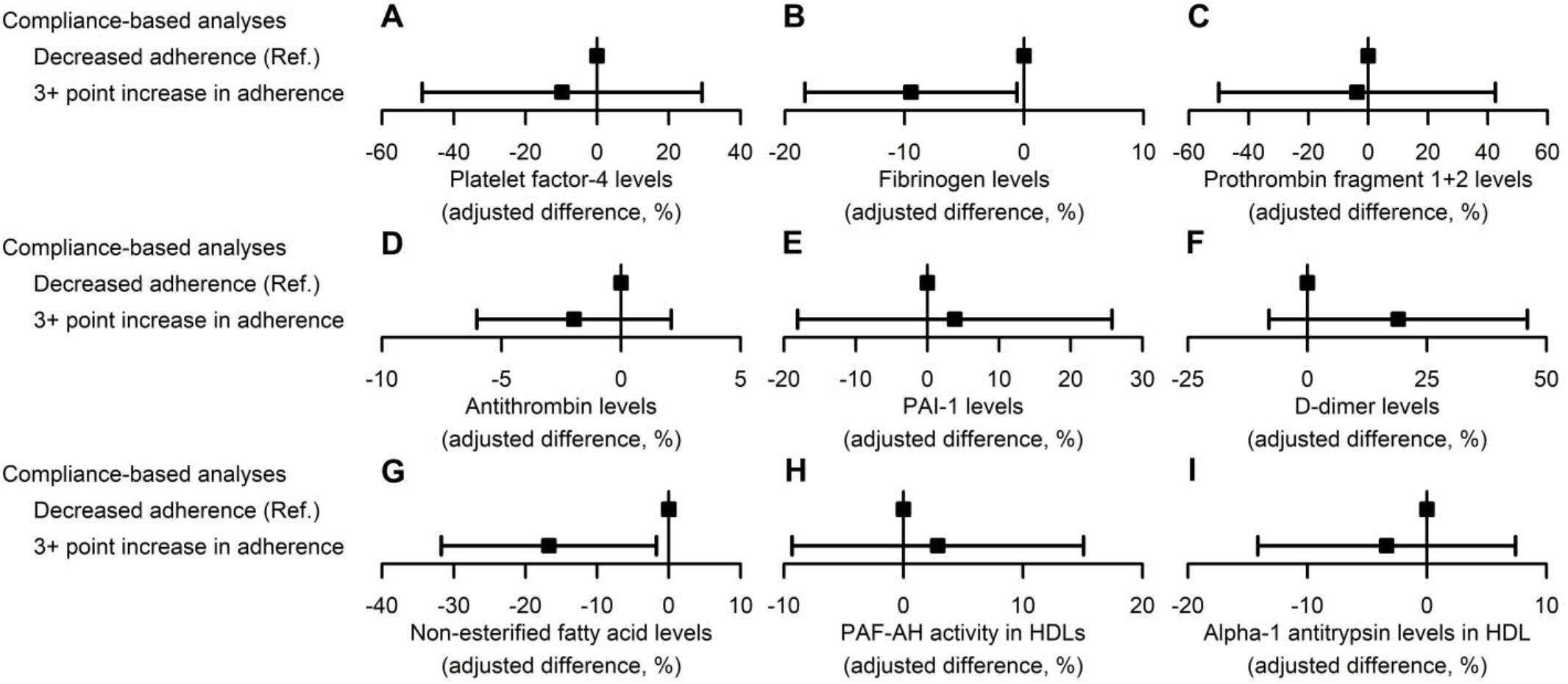
Associations between substantial increments in MedDiet adherence after 1 year of intervention (≥3 score points) with 1-year changes in atherothrombosis biomarkers (relative to 1-year adherence decreases). Results are shown as adjusted coefficients (percentage changes relative to baseline values) with 95% confidence intervals. *HDL*: high-density lipoprotein; *PAF*-*AH*: platelet activating factor acetylhydrolase; *PAI-1*: plasminogen activator inhibitor 1.

Exact coefficients of all analyses in the whole study population are available in **Supplemental Table 5**.

## DISCUSSION

Our results show that following a MedDiet improves several atherothrombosis biomarkers in high cardiovascular risk individuals.

Atherothrombosis is strongly affected by inflammation and oxidative stress^[25,26]^. MedDiet is a lifestyle intervention known to enhance these risk factors^[4,27]^, possibly explaining its protective effects on thrombosis as well. In our data, following the MedDiet-VOO intervention improved HDL antithrombotic properties (and, under certain circumstances, antithrombin and D-dimer levels as well), whilst the MedDiet-Nuts decreased NEFA levels. We previously reported that the MedDiet-VOO intervention enhanced HDL functionality in the PREDIMED Study^[21]^. However, these findings extend this protection to HDL antithrombotic functions. The MedDiet antioxidant protection on HDL components could contribute to explaining the reported improved activity of HDL protective enzymes (due to a greater preservation of their non-oxidized structure)^[28]^. In addition, the MedDiet capacity to decrease plasma levels of inflammatory biomarkers may help explain the reduction in the concentrations of pro-inflammatory molecules bound to HDL particles^[15]^. This decrease in low-grade inflammation could also contribute to explaining the enhancement of other pro-thrombotic signals (the decrease in D-dimer levels after the MedDiet intervention among users of antithrombotic drugs agrees with previous cross-sectional evidence^[17]^). In relation to NEFAs, they promote endothelial stress and platelet aggregation and are considered an emergent pro-thrombotic risk factor in high cardiovascular risk states such as obesity and hypertriglyceridemia^[13]^. As hypothetical mechanisms, polyunsaturated fatty acids (whose intake was particularly increased in the MedDiet-Nuts group) bind to GPR120 receptors in adipose tissue and decrease lipolysis, decreasing the circulating levels of free fatty acids^[29]^. In addition, MedDiet is a fiber-rich dietary pattern, and the intestinal metabolism of fiber leads to blood peaks of short-chain fatty acids (known to be able to bind other GPR family receptors with a similar anti-lipolytic effect^[30]^). Finally, substantial increments in MedDiet adherence were also associated with lower fibrinogen concentrations (as well as with less NEFA levels). Fibrinogen is a biomarker known to be a connection point for inflammation and thrombosis^[9]^. Decreased fibrinogen concentrations in our findings could also be partially explained by the MedDiet anti-inflammatory capacity^[4]^ and agree with previous cross-sectional evidence in humans^[18]^.

Our study has limitations. First, our analyses were not originally included in the study protocol and, therefore, should be considered as exploratory findings. Second, study volunteers were elderly people at high cardiovascular risk, which does not allow the extrapolation of our findings to other populations. Third, we have only been able to report moderate changes or associations. However, this effect magnitude was expected because our work was based on modest real-life dietary modifications and the control diet was already a well-known healthy dietary pattern. Finally, data on MedDiet adherence (and other covariates such as physical activity and ethanol intake) were self-reported and could be somewhat imprecise. However, the main strengths of our study were its large sample size (*N*=358), randomized design, long-term duration (1 year), and strict quality control for laboratory biomarkers.

In conclusion, 1 year of intervention with MedDiet enhanced HDL antithrombotic properties and decreased NEFA levels in high cardiovascular risk individuals (and at a second level of relevance, increased antithrombin and reduced D-dimer concentrations in certain study subpopulations). As observed in the secondary, observational analyses, substantial increments in MedDiet adherence were associated with decreases in fibrinogen and NEFA levels. As far as we know, this is the largest, most comprehensive, hypothesis-based analysis to date of the effects of a healthy dietary pattern on atherothrombosis indicators in high cardiovascular risk individuals. Our results support the improvement of thrombosis status after following a MedDiet. Further studies are needed to confirm whether these improvements mediate the reported cardioprotective benefits of such lifestyle modifications.

## Data Availability

Considering that we do not have the explicit written consent of the study volunteers to yield their deidentified data at the conclusion of the study, individual participant data cannot be shared. The study protocol is available in the main publication of the study1 and in the study website (http://www.predimed.es/uploads/8/0/5/1/8051451/_1estr_protocol_olf.pdf), and a summary of the dietary intervention is also available in Supplemental Methods.

## AUTHOR CONTRIBUTIONS

A.H. and R.C. acquired data. A.H. analyzed and interpreted data. X.P., M.Fitó, M.A.M.-G., D.C., J.S.-S., J.L., E.G.-G., F.A., M.Fiol, L.S.-M., X.P., E.R., and R.E. conceived the clinical trial concept and design. A.H., O.C., A.T.-R., X.P., and R.E. obtained funding. A.H. drafted the manuscript. O.C., A.T.-R., X.P., M.Fitó, R.C., M.A.M.-G., D.C., J.S.-S., J.L., E.G.-G., F.A., M.Fiol, L.S.-M., X.P., E.R., and R.E. revised the content of the manuscript and approved its final version. A.H. is the guarantor of this work and, as such, had full access to all of the data in the study and takes responsibility for the integrity of the data and the accuracy of the results.

## ACKNOWLEDGEMENTS

This work was supported by grants of Instituto de Salud Carlos III [OBN17PI02, PIE14/00045_INFLAMES, CB06/03/0019, CB06/03/0028, and CD17/00122 (A.H.)], and Agència de Gestió d’Ajuts Universitaris i de Recerca (2017 SGR 222).

Authors wish to thank Daniel Muñoz-Aguayo, Gemma Blanchart, and Sònia Gaixas for their technical assistance, and Stephanie Lonsdale for revising the English text. CIBER de Fisiopatología de la Obesidad y Nutrición (CIBEROBN) is an initiative of the Instituto de Salud Carlos III, Madrid, Spain.

## Abbreviations

MedDiet: Traditional Mediterranean Diet
MedDiet-Nuts: Traditional Mediterranean Diet enriched in nuts
MedDiet-VOO: Traditional Mediterranean Diet enriched in virgin olive oil
NEFA: non-esterified fatty acids
PAF-AH: platelet activating factor-acetylhydrolase
PAI-1: plasminogen activator inhibitor-1

